# Dopaminergic modulation of local non-oscillatory activity and global-network properties in Parkinson’s disease: an EEG study

**DOI:** 10.1101/2021.12.17.21267975

**Authors:** Juanli Zhang, Arno Villringer, Vadim V. Nikulin

## Abstract

Dopaminergic medication for Parkinson’s disease (PD) modulates neuronal oscillations and functional connectivity across the basal ganglia-thalamic-cortical circuit. However, the non-oscillatory component of the neuronal activity, potentially indicating a state of excitation/inhibition balance, has not yet been investigated and previous studies have shown inconsistent changes of cortico-cortical connectivity as a response to dopaminergic medication. To further elucidate changes of regional non-oscillatory component of the neuronal power spectra, functional connectivity, and to determine which aspects of network organization obtained with graph theory respond to dopaminergic medication, we analyzed a resting-state EEG (Electroencephalogram) dataset including 15 PD patients during OFF and ON medication conditions. We found that the spectral slope, typically used to quantify the broadband non-oscillatory component of power spectra, steepened particularly in the left central region in the ON compared to OFF condition. In addition, using lagged coherence as a functional connectivity measure, we found that the functional connectivity in the beta frequency range between centro-parietal and frontal regions was enhanced in the ON compared to the OFF condition. After applying graph theory analysis, we observed that at the lower level of topology the node degree was increased, particularly in the centro-parietal area. Yet, results showed no significant difference in global topological organization between the two conditions: either in global efficiency or clustering coefficient for measuring global and local integration, respectively. Interestingly, we found a close association between local/global spectral slope and functional network global efficiency in the OFF condition, suggesting a crucial role of local non-oscillatory dynamics in forming the functional global integration which characterizes PD. These results provide further evidence and a more complete picture for the engagement of multiple cortical regions at various levels in response to dopaminergic medication in PD.

## 1. Introduction

Parkinson’s disease (PD) is the second most common neural degenerative disorder characterized by massive degeneration of dopaminergic neurons in the nigrostriatal dopamine system (Olanow et al., 2009). It has been increasingly recognized that PD is accompanied by functional disturbances both at subcortical and cortical levels (Boon et al., 2019; Braak et al., 2003). Clinically, dopamine loss is managed via dopaminergic therapy (DT). The dopaminergic system has been shown to have considerable and widespread modulatory influences on many brain structures including the cortex (Steiner & Kitai, 2001). While dopamine replacement therapy is efficient for improving the motor symptoms, the neural mechanisms of dopaminergic medication are not yet fully understood (Schapira, 2005).

In PD, it has been repeatedly reported that it is characterized by abnormal oscillatory synchrony in the basal ganglia-thalamus-cortical (BGTC) network in the beta frequency band (13–30Hz) that could be modulated by dopaminergic medications and deep brain stimulation (DBS) (Brown, 2003; De Hemptinne et al., 2015; Kühn et al., 2009; Müller & Robinson, 2018; Wingeier et al., 2006). In the frequency domain, electrophysiological brain signals typically consist of a power-law 1/f component and periodic oscillatory activities. While a majority of studies have so far been dedicated to the oscillatory activity, increasing evidence shows that non-oscillatory (aperiodic) activity also provides information about the intricate neuronal dynamics unfolding at different temporal scales (He et al., 2010; Voytek et al., 2015). A broadband aperiodic component of the spectrum is often represented by the slope of the fitted line in log-log space (known as spectral slope). The changes in spectral slope have been associated with neural development, healthy aging, and performance in working memory tasks (Donoghue et al., 2020; Voytek et al., 2015). In addition, previous studies have reported that it is altered in different pathologies, such as schizophrenia (Molina et al., 2020; Peterson et al., 2017) and ADHD (Attention deficit/hyperactivity disorder) (Robertson et al., 2019). Importantly, it has also been demonstrated that the spectral slope is a potential indicator of the local excitation/inhibition balance (R. Gao et al., 2017; Colombo et al., 2019). In addition, TMS (transcranial magnetic stimulation) studies, which can directly probe the changes in excitation and inhibition, have shown that PD is accompanied by changes in cortical excitability (Cantello, 2002; Hanajima et al., 1996; Ridding et al., 1995). Thus, it would be important to test whether and how this measure is altered in PD, in particular with dopaminergic medication.

While regional changes could provide comprehensive understanding of the underlying local circuitry, the brain rather functions as a distributed network. Functional connectivity analysis allows us to understand how distinct regions interact, and graph-theory based approach enables a macroscopic perspective of brain connections on the regional and whole-brain network level. Many previous studies showed that network architecture is related to brain function or dysfunction (Bassett & Bullmore, 2009; Bullmore & Sporns, 2009). Using resting state fMRI (functional magnetic resonance imaging), it has been intensively investigated how dopaminergic medication modulates brain functional connectivity in the BGTC network (Tahmasian et al., 2015). The most consistent finding across different rs-fMRI studies revealed decreased connectivity within the posterior putamen in PD (Tessitore et al., 2019), and that its cortical projections are modulated by dopaminergic medication (Herz et al., 2014). To date, few fMRI studies have adopted graph theoretical approach in PD, and the reported findings have been inconsistent. Specifically, compared to healthy controls, PD patients showed lower global efficiency (Sang et al., 2015), while no abnormalities in topographical property at the global level were observed in PD (Berman et al., 2016; Hou et al., 2018; Ruan et al., 2020). Both increase (Sang et al., 2015) and decrease (Hou et al., 2018) in nodal centrality have been observed in PD compared to healthy controls. In addition, it was found that levodopa administration significantly decreased local efficiency of the network (Berman et al., 2016), and conversely resulted in an increase in eigenvector centrality of cerebellum and brainstem in PD (Jech et al., 2013).

As for the EEG/MEG (Electro- and Magnetoencephalography) studies, compared to healthy controls, increased cortico-cortical functional connectivity in PD has been found primarily in alpha and beta frequency ranges, and cortico-cortical coherence was linked to the severity of the clinical symptoms (Bosboom et al., 2009; George et al., 2013; Miller et al., 2019; Silberstein et al., 2005; D. Stoffers et al., 2007; Diederick Stoffers et al., 2008). Dopaminergic medication induced changes in cortical synchronization have also been investigated by computing pair-wise coherence across the entire montage using multi-channel EEG/MEG. However, both reduction of functional connectivity after dopamine medication (George et al., 2013; Heinrichs-Graham et al., 2014; Silberstein et al., 2005) and the absence of connectivity modulation were previously reported (Miller et al., 2019). Very recently, using advanced modeling analysis, in response to dopaminergic medication, increased cortico-cortical synchronization in beta band has been detected by taking into account the contribution from other sub-networks (Sharma et al., 2021). To capture the changes across the whole cortex, through the application of graph theoretical measures in EEG/MEG, previous studies have demonstrated abnormalities in topographical organizations of functional network in PD compared to healthy controls, suggesting that the interactions between cortical areas become abnormal and contribute to PD symptoms at various stages (Utianski et al., 2016). Furthermore, the alterations in network attributes were linked to both motor and cognitive dysfunctions (Boon et al., 2017; Olde Dubbelink et al., 2014). However, how the topological organization of the cortical functional network changes after dopaminergic administration remains rather elusive. To address this issue, we applied graph theory-based network analysis to investigate further changes in cortical connectivity in patients with PD after the administration of dopaminergic medication.

To further characterize the regional and functional network changes due to dopaminergic medication, we address the following questions. Regarding local properties: 1) How does the aperiodic property of the electrophysiological brain signal change in response to dopaminergic medication administration? With respect to cross-area interactions: 2) What is the effect of dopaminergic medication on functional connectivity? 3) Does dopaminergic medication induce alterations in the lower and/or higher level of the network architectures? 4) Do local changes in non-oscillatory component of neural activity influence functional network topology/organization? To answer these questions, we analyzed a publicly available dataset including EEG data of PD patients from ON and OFF dopaminergic medication conditions (George et al., 2013; Rockhill et al., 2020).

## 2. Methods and Materials

### 2.1. Participants

The data analyzed in this study is open-source data (George et al., 2013; Jackson et al., 2019; Swann et al., 2015). This dataset includes resting state EEG data with a duration of around 3 min. Data were collected from 15 PD patients (8 female, average age = 63.2 ± 8.2 years, mild to moderate disease with average disease duration of 4.5 ± 3.5 years) during OFF and ON dopaminergic medication sessions. All participants were right-handed and provided written consent in accordance with the Institutional Review Board of the University of California, San Diego and the Declaration of Helsinki. For more information you may refer to George et al. (2013).

### 2.2. Data collection

EEG of patients with PD were recorded on two different days for ON and OFF medication sessions which were counterbalanced across subjects. For the OFF medication session, patients were requested to withdraw from their medication at least 12 h prior to the EEG recording. For the ON medication session, subjects took their medication as usual. A 32-channel EEG cap with BioSemi ActiveTwo system was used to acquire the EEG data with a sampling rate of 512 Hz. Two additional electrodes were placed over the left and right mastoids used for reference. During the EEG recording, participants were instructed to sit comfortably and fixate on a cross presented on the screen. Each recording session lasted at least 3 minutes. In addition, participants completed a few clinical assessments which were previously reported in George et al. (2013). In this study, we did not include the clinical scores of patients as the authors of the original paper mentioned some uncertainty about these scores.

### 2.3. Data pre-processing

EEG data were analyzed using EEGLAB (version 14.1.2; Delorme & Makeig, 2004) and FieldTrip toolboxes, together with customized scripts in Matlab (The MathWorks Inc, Natick, Massachusetts, USA). First, a high-pass filter at 1 Hz was applied to remove low frequency drifts (two-way FIR filter, order = 1536, eegfilt.m from EEGLab). Subsequently, independent component analysis (ICA – Infomax algorithm implemented in EEGLab) was used to remove artifactual sources of cardiographic components, eye movements and blinks, and muscle activity in the data. Further, channels with inadequate quality were rejected by visually inspecting whether their spectra demonstrated residual EMG at higher frequency ranges (on average 5.4±3.1 for OFF and 5.2±2.8 for ON, no difference between conditions (p = 0.6606)). Bad channels were interpolated with neighboring electrodes using a method of spherical splines (EEGLab function ‘*eeg_interp’*). Next, data were examined visually for the presence of residual artifacts and segments contaminated by gross artifacts and these events were marked and then excluded from further analysis (on average 172.5±22.7s in OFF and 165.5±33.6s in the ON condition remained, no difference in the number of rejected data points (p = 0.3591)). Subsequently, data were re-referenced to the common average.

## 3. Data analysis

### 3.1. Power spectral density (PSD)

Power spectral density (PSD) was calculated using the function ‘*pwelch’* in MATLAB, with a Hamming window of 512 samples (i.e., 1 second) and a 50% overlap. The average PSD for the beta band was obtained by averaging the spectral density in the beta frequency range (13–30 Hz).

### 3.2. PSD Slope

To reduce contamination from high frequency non-neuronal noise, we estimated the slope of the PSD in a frequency range of 2–45Hz. A three-step robust regression method was used to estimate the slope based on the computed PSD. This method was proposed and applied by Colombo et al. (2019). First, a least-squares linear line was fitted to the raw PSD using the function ‘*robustfit*’ in MATLAB in the log frequency-log PSD space. Second, frequency points with larger than 1 median absolute deviations of the PSD residuals were identified as oscillatory peaks. Continuous frequency bins surrounding these peak frequencies were considered as the base of the oscillatory peaks and were also excluded for the further step. Last, a second least-squares fit was performed on the rest of the frequency ranges. We took the slope (with the sign) of the second fitted line as the final spectral slope of the PSD. Thus, a more negative slope demonstrates a steeper decay, while a less negative slope represents a flatter one. One advantage of this method is that it considers the potential bias resulting from linearly spaced frequency bins being estimated with a logarithmic scale. Therefore, before the regression procedure, the PSD curve was up-sampled with logarithmically distributed frequency bins. For more details, please refer to the study by Colombo et al. (2019).

### 3.3. Functional network analysis

A network is constructed by a collection of nodes and links between pairs of nodes. In this study, we defined each node as a brain region approximately represented by each channel, while links represent the connectivity between pairs of channels. Functional connectivity between the brain areas was determined by computing the lagged coherence which accounts for the volume conduction issue. Each network can be represented by a symmetrical 32*32 adjacency matrix.

#### 3.3.1. Functional connectivity

Functional connectivity (FC) measure was quantified by the lagged coherence between all the channel pairs in a frequency range of 1 – 35 Hz with resolution of 1 Hz. This metric quantifies the strength of phase coupling between two signals by eliminating the effects of volume conduction (Pascual-Marqui, 2007; Pascual-Marqui et al., 2011), and it has been shown to be even more suitable than phase lag index for the application of connectivity estimation when using EEG and MEG (Hindriks, 2021). Its value ranges between [0, 1]: “0” stands for no coupling, and “1” represents perfect coupling. This measure has been utilized in earlier EEG studies (Milz et al., 2014; Vecchio et al., 2021). Functional connectivity in an oscillatory frequency band was acquired by averaging the FC values over the respective frequency range (for instance beta band FC was obtained by averaging the FC values over 13–30Hz and 8–12Hz for the alpha band).

#### 3.3.2. Network measure

We estimated the brain network metrics based on the scalp sensor-based EEG connectivity matrix. Although often performed in source space, due to a small number of channels (Lantz et al., 2003) we did it rather in sensor space similar to previous studies (Chai et al., 2019; Mitsis et al., 2020; Smith et al., 2021; Stam et al., 2007; Sun et al., 2019; Zeng et al., 2015). In the discussion, we mention and discuss limitations associated with the estimation of graph metrics in sensor space.

##### 3.3.2.1. Node degree

Node degree estimates the number of edges connected to each node. To estimate the importance of each node (each channel in our case), node degree centrality weighted by edge importance (the connection is stronger, edge weights are larger) was utilized for this purpose. Specifically, we used the function ‘*Centrality’* implemented in Matlab for this measure (parameter “*importance*” specified by edge weights).

##### 3.3.2.2. Graph theory based complex network measures Overall functional connectivity

For each individual functional connectivity matrix, the overall functional connectivity was obtained by averaging all the connectivity values across all the pairs of the connection in a matrix.

##### Proportional thresholding

Proportional thresholding is a commonly applied approach to remove connections with lower strength and to obtain a sparse connectivity matrix for computing the network properties based on graph theory. Here, we applied a proportional threshold to keep a consistent density of the connections across individuals (Bassett & Bullmore, 2009; van den Heuvel et al., 2017). If a proportional threshold (PT%) is applied to a functional network, all the strongest PT% of the connections are preserved and set to 1; the other connections are set to 0. As suggested by (Rubinov & Sporns, 2010), networks should be ideally characterized and show consistent patterns across a broad range of thresholds. These threshold values are often determined differently across studies. Therefore, in this study we examined a wide range of thresholds ranging from 36% to 4% (resulting in networks with around 20 ∼ 200 links) in steps of 2%, similar to a previous study (van den Heuvel et al., 2017).

##### Graph metrics

Various measures characterize a network’s structure. Two fundamental ones are included here: clustering coefficient and global efficiency. These two basic graph metrics were computed as implemented in the Brain Connectivity Toolbox (Rubinov & Sporns, 2010). Clustering coefficient is a commonly used measure to quantify the functional network segregation. It is defined as the fraction of triangles (ratio of the present and total possible number of connected triangles) around an individual node and is equivalent to the fraction of a node’s neighbors that are neighbors of each other (Watts & Strogatz, 1998). The clustering coefficient of a network CC is the average clustering coefficient across all the nodes in the network. It reflects the prevalence of clustered connectivity around individual nodes (Rubinov & Sporns, 2010): the larger the CC, the greater the degree of functional segregation.

The other metric, global efficiency (GE), was used to quantify the functional network integration. This is based on a basis measure – shortest characteristic path length. Paths are sequences of distinct nodes and links, with shortest paths between two nodes defined as the path with the fewest edges in a network (the sum of the number of its constituent edges is minimized). Global efficiency for a network, obtained by the average inverse shortest path length between all the pairs, is a measure of functional network integration: the larger the GE, the greater the degree of global integration. All these measures were computed with an open source Matlab toolbox (Rubinov & Sporns, 2010; http://www.brain-connectivity-toolbox.net).

### 3.4. Statistical tests

Non-parametric Wilcoxon signed rank test was performed for the comparisons of measures in PD OFF and ON states. Spearman’s correlation coefficients were calculated to estimate the relations between different measures. We applied the false discovery rate (FDR) procedure (Yoav Benjamini & Yosef Hochberg, 1995) to correct for multiple tests (correlation calculation) across channels. Significance is reported when FDR-corrected p-values are below 0.05.

To account for multiple comparisons of metrics across all channels, we performed a channel space cluster-based permutation test using the ‘*Monte Carlo*’ method, as implemented in FieldTrip (Oostenveld et al., 2011). At sample level (each channel in this case), a dependent T-test was utilized to estimate the effect. 1000 randomizations were performed across groups (ON and OFF conditions) and for each permutation. Additionally, the single sample t-values are thresholded at the 95-th quantile, and cluster-level statistics (sum of t-values within each cluster) were computed and the largest cluster statistic was taken to build a null distribution. We then compared the observed cluster-level statistic from the empirical data against the null distribution derived from the permutation procedure. P values below 0.05 (two tailed) were considered significant. A positive or negative cluster demonstrates a significant difference between two conditions (OFF > ON) or (OFF < ON).

## 4. Results

### 4.1. Spatial specificity and effects of medication on spectral slope

The grand mean of PSD averaged from all channels across subjects in each group is shown in Fig. 1A. One can observe that the PSD decay in PD OFF was shallower compared to the PSD decay in PD in the ON condition. The spectral slope was computed for each channel and each subject. Figure 1B shows the topography of the grand mean of the spectral slope across all subjects within each group (upper panel for OFF and lower panel for ON condition). As shown in Figure 1B, for both groups, spectral slopes were more negative (steeper slopes) along the fronto-central-parietal midline of the brain and flatter in the other regions. In general, the ON condition was characterized by a more negative slope than that in the OFF condition.

**Figure 1.**
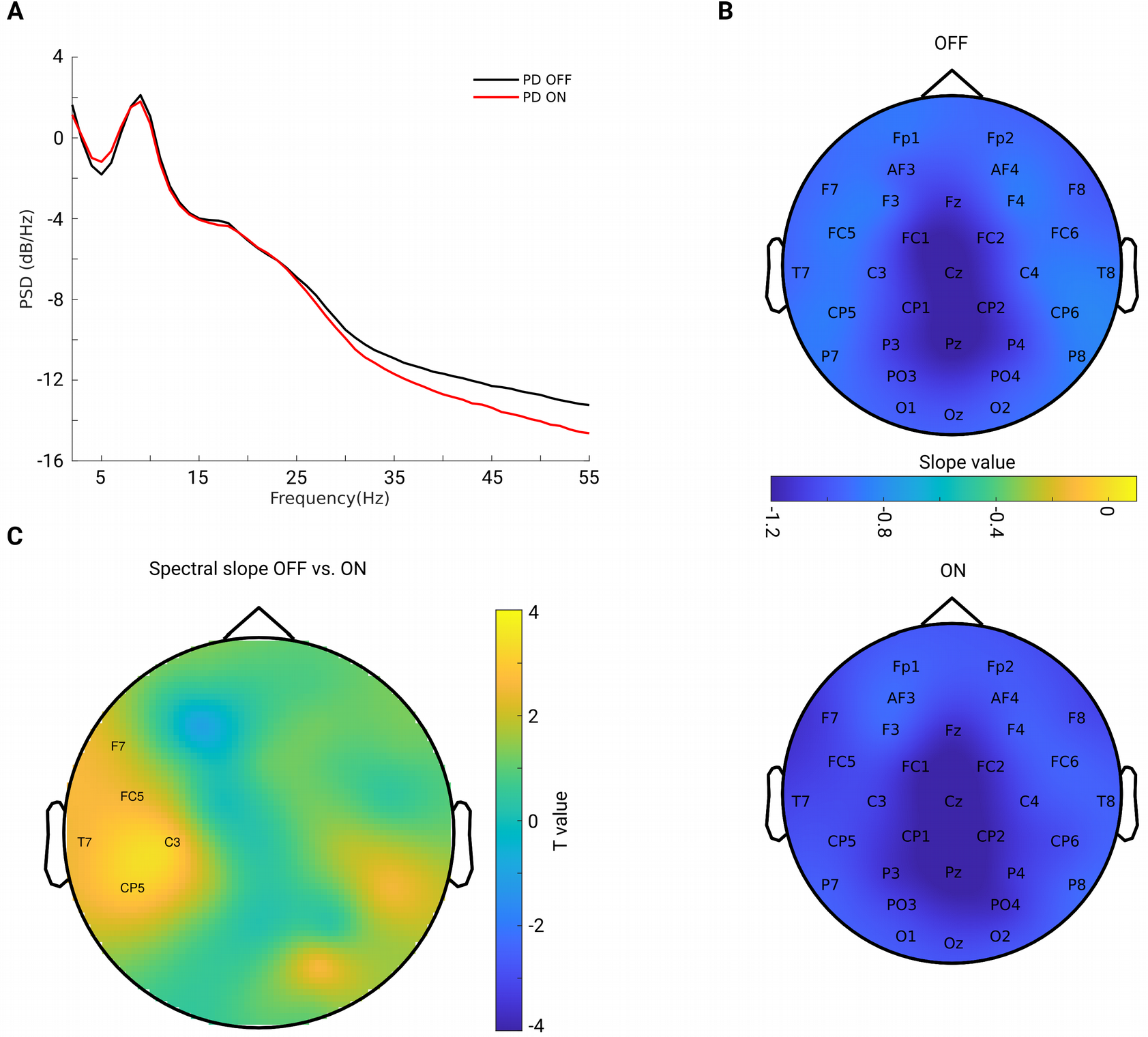
A. Grand mean of PSD across all channels and subjects within each group (OFF in black and ON in red). B. Grand mean spatial distribution of spectral slope estimated from power spectra over 2–45Hz across subjects within each group (upper panel for OFF, lower panel for ON condition). Color bar indicates the slope value. C. Spatial-difference pattern of spectral slope between OFF and ON (OFF-ON) condition (cluster-based permutation test, p = 0.0220). Significant positive clusters are labeled. Color bar indicates the statistical T-value.

We investigated the difference between the two conditions for all channels. As described in the Methods section, we applied a non-parametric cluster-based permutation test to correct for multiple comparisons in the channel space. When comparing slope values in PD OFF with those of PD ON, a significant positive cluster (p = 0.0220) indicated an increased slope (flatter) in PD OFF. This difference demonstrated a lateralized pattern covering mostly left central region (Fig. 1C).

### 4.2. No beta power difference between conditions before and after correcting for the slope effect

Previous studies have demonstrated inconsistent changes in cortical beta power: an increase of beta power after dopaminergic medication (Melgari et al., 2014) and insignificant cortical beta power changes after dopaminergic therapy in PD (George et al., 2013; Miller et al., 2019). Since we showed that the background slope was significantly modulated by dopaminergic medication (significantly steepened by the medication), we assumed that insignificant beta power reports might partly be attributed to the overall broadband slope changes. To test this assumption, we first applied a traditional approach to estimate the beta band power by averaging the PSD values in the beta frequency range (13 – 30 Hz) based on the raw PSD. We computed the mean PSD value in the beta frequency range (13 – 30 Hz) for each channel and each subject in each group. Cluster-based permutation tests in channel space showed no significant difference in beta power between conditions (Fig. 2A). Next, to address whether this finding might be due to a flattened background spectral slope (as observed in the PD OFF versus ON comparison) on the top of which oscillations were present, we used a second approach controlling for the spectral slope to estimate beta-oscillation power. In line with a previous study (Donoghue et al., 2020), we subtracted the fitted straight line in log-log space of the PSD spectra and calculated the mean PSD values in a given frequency range (13–30Hz for beta) on the residuals of the PSD spectra for each channel and subject. Figure 2B shows the grand mean of the residuals of the PSD across all channels after accounting for spectral slope. By averaging the PSD values in the same frequency range of 13–30Hz, beta band power for each channel and each subject was re-calculated. Cluster-based permutation tests identified two non-significant negative clusters (OFF-ON) (p = 0.0739, 0.0939), mainly localized in bilateral centro-parietal regions (CP5, CP1 and C4, CP6, Fig. 2C). This demonstrates that even after accounting for the background slope effect, there were no significant beta power changes between the two medication conditions.

**Figure 2.**
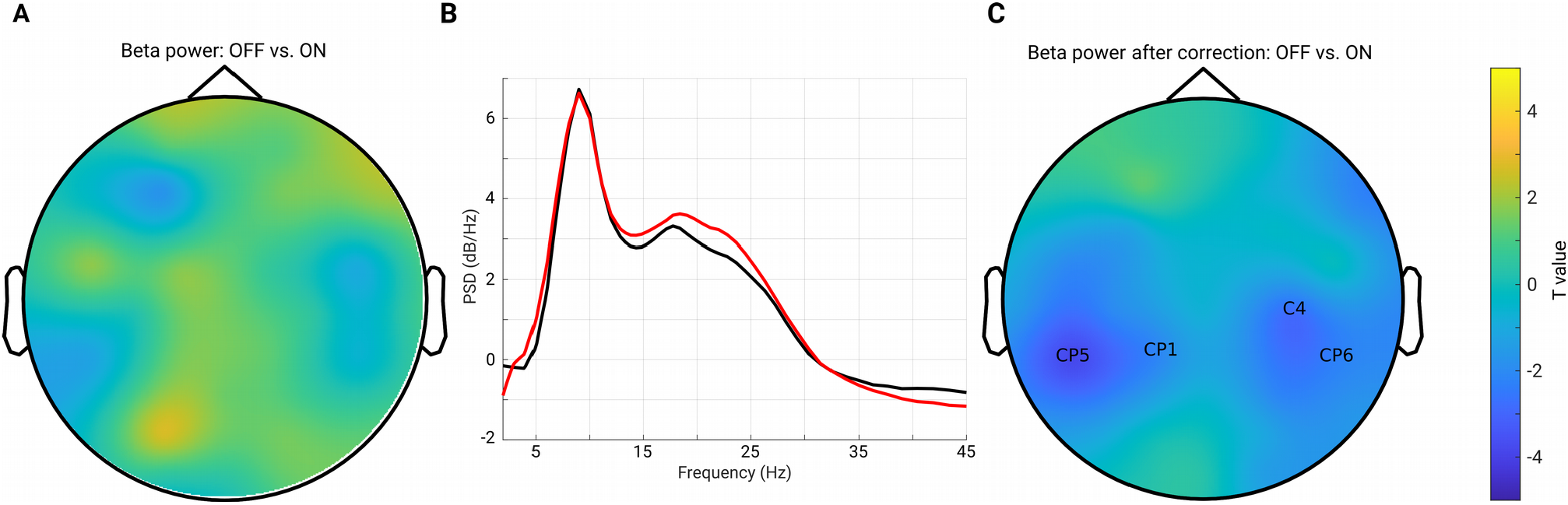
A. Topography for the comparison of beta band power between PD OFF and ON conditions estimated from the raw power spectra. No significant cluster was detected for the comparison. B. Mean of the residuals of PSD (across all channels and subjects within each group, OFF in black and ON in red) after subtracting the spectral slope. Oscillatory peaks are present in alpha and beta bands in both groups. C. Topography for the comparison of oscillatory beta band power between PD OFF versus ON conditions after accounting for the background spectral slope. Two negative clusters were identified as shown by the labels, but none of them reached significance (p = 0.0739, 0.0939). Color bar indicates the statistical T-value.

### 4.3. Functional connectivity in beta band is increased after medication

The lagged coherence averaged over the frequency of interest (such as alpha or beta) was taken as a connectivity measure in this frequency band. To investigate whether medication could result in changes in functional connectivity in oscillatory frequency band across the whole brain (neighboring areas and remote regions), we applied a seed-based connectivity comparison approach. This means that the connectivity was calculated between a given electrode (seed) and all other electrodes for each subject. Then, whole-head connectivity was compared between conditions using a cluster-based permutation test to account for multiple comparisons. First, we predominantly focused on the sensorimotor seed-based connectivity changes, which typically include C3 and C4 electrodes (Miller et al., 2019; Swann et al., 2015). The upper panel of Figure 3A depicts the FC between C3 and one of the representative channels from the parietal region (Pz) along a wide frequency range (1–35Hz). One can observe clear peaks around the alpha and beta frequency bands for both the ON and OFF conditions. Next, we averaged the connectivity values in the beta frequency range (13 – 30 Hz) as a measure of beta band functional connectivity. As described above, C3 seed-based beta band connectivity was compared between medication conditions. A negative cluster localized in the parieto-occipital region (OFF < ON, p = 0.007) was identified as shown in the upper panel of Figure 3B, demonstrating a lower connectivity between C3 and parieto-occipital regions in the OFF compared to the ON conditions. However, there was no significant difference in the comparison of C4 seed-based connectivity between conditions. Furthermore, to investigate whether the frontal region showed altered synchronization with other regions, we chose one of the representative channels in the frontal area (Fz, which is typically within the cluster of electrodes near the supplementary motor area (Casarotto et al., 2019)) and performed the same analysis as for electrode C3. As shown in the lower panel of Figure 3A, there were obvious peaks in the broad oscillatory frequency range (alpha and beta) for both conditions. The lower panel of Figure 3B shows the topographical pattern for the comparison between OFF and ON conditions, and a significant negative cluster (p = 0.0250) localized primarily in the parietal region. This demonstrated that the synchronization between Fz and parietal regions in the beta band was significantly enhanced in the ON compared to OFF condition in PD. Finally, we performed the same analysis for the other channels to demonstrate whole-head comparisons in a head-in-head plot (Fig. 3C). As in C3 and Fz seed-based connectivity comparisons, the other channels in seed-based connectivity also showed significant increase in ON compared to OFF conditions. Significant clusters (p < 0.05) are marked by warm color. In general, the topographies showed significant alterations in synchronization between frontal, central and parieto-occipital regions. In addition, due to presence of peaks of the functional connectivity in the alpha band, we used the same approach to explore the functional connectivity changes in alpha band (8-12Hz). Yet, there was no significant cluster detected for all the possible seeds when comparing the two conditions. Due to our predominant interest in the beta frequency range and pronounced effects observed in this frequency band, in the rest of the study we focus on the measures from the beta band.

**Figure 3.**
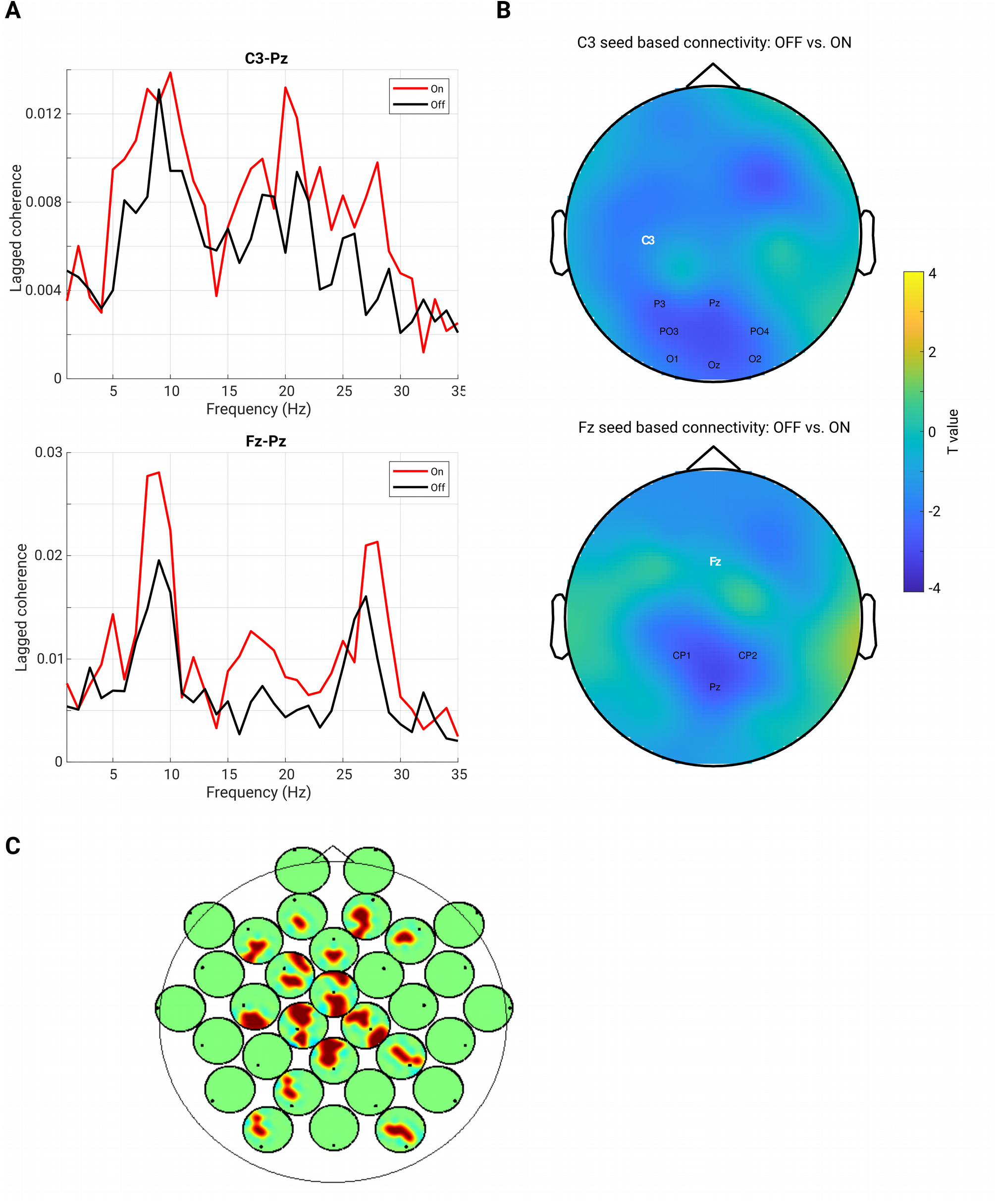
A. Lagged coherence plot over a frequency range of 1 - 35 Hz. Upper panel shows the mean connectivity (measured by lagged coherence) estimated from C3 and Pz, while the lower panel shows the connectivity estimated from Fz and Pz, across the subjects within each group (OFF in black and ON in red). B. Upper panel: Topography for C3 seed-based connectivity (lagged coherence in beta band) comparison between OFF vs. ON condition (channel-space cluster-based permutation test). The significant cluster is highlighted by the labels in black, while the seed channel C3 is marked in white. Lower panel: the same analysis performed for the seed channel Fz, and a significant negative cluster (OFF < ON) was detected (p = 0.0250). Color bar indicates the statistical T-value. C. Head-in-head plot for the seed-based connectivity (lagged coherence in beta band) comparison for all channels. At each channel, the head plot shows the topography for comparison of connectivity between this channel and all other channels using cluster-based permutation test. Only the significant clusters (p < 0.05) are shown by warm color.

### 4.4. Node degree in centro-parietal region in beta band is increased after medication

Next, we tested whether the local level of a network feature, namely the node degree, was modulated by the medication effect. For this purpose, we calculated the node degree (from the connectivity in the beta band) for each channel and each subject. Figure 4A shows the topographical maps of the grand mean of the node degree across subjects within each group. As can be seen from Figure 4A, both groups showed a spatial specificity regarding the degree distribution (left for OFF and right for ON conditions): a higher level of the node degree in central areas than in other regions. This demonstrates that the central region might, in general, interact more with other regions in the whole brain network. Next, we compared the node degree between conditions for all channels using a cluster-based permutation test. Figure 4B shows the spatial difference pattern – a significant negative cluster was detected (p = 0.0140, OFF vs. ON, shown by labels) mainly in the centro-parietal region, suggesting that medication modulated the node degree of the beta band functional network in a way that the connectivity of the centro-parietal region became more pronounced in the whole network. Thus, this analysis further confirmed our findings obtained from seed-based connectivity analyses, revealing that synchronization was up-regulated by medication specifically between the centro-parietal region and other regions.

**Figure 4.**
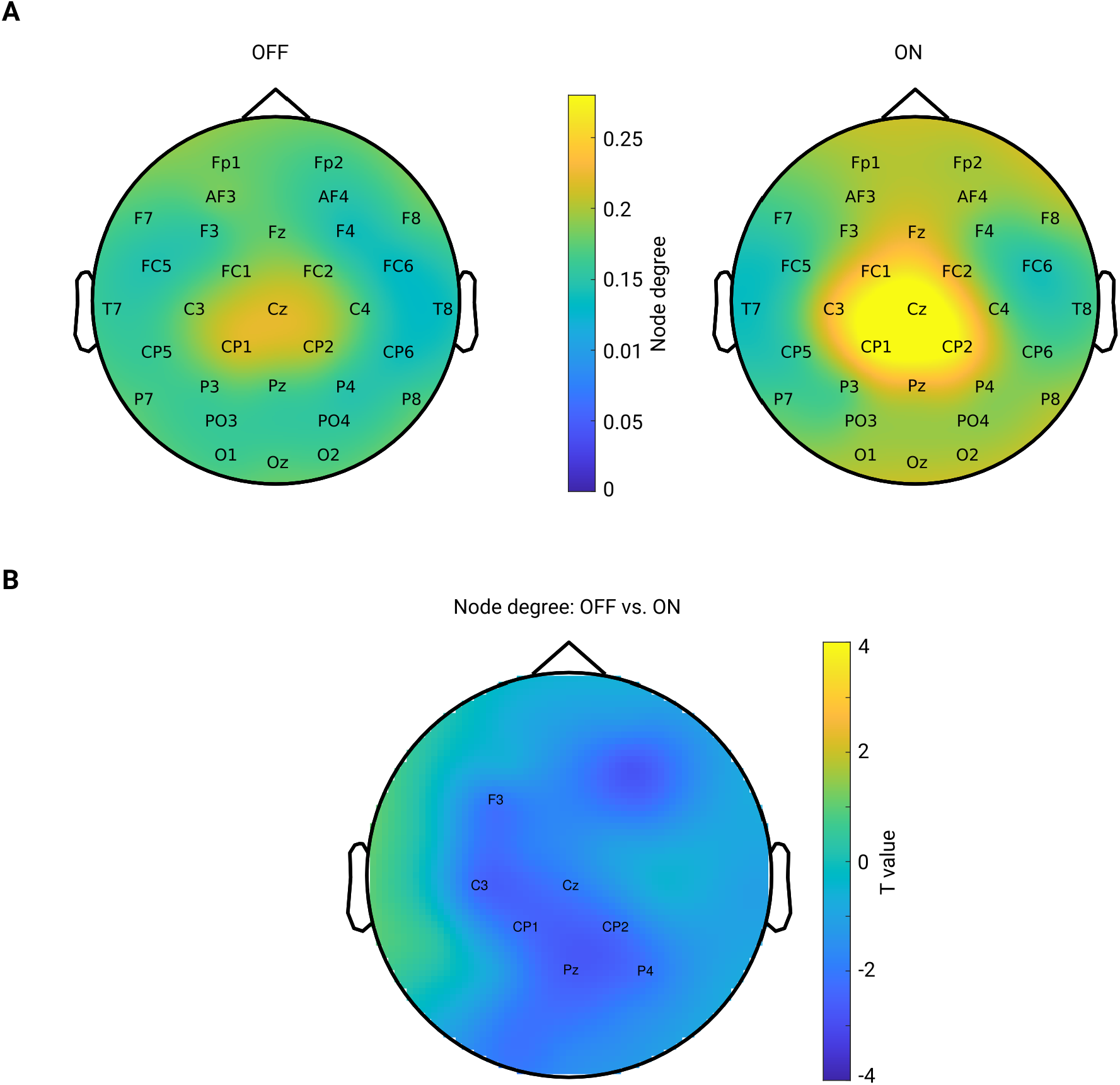
A. Mean spatial distribution of node degree calculated from the beta band functional connectivity for each group: Left for OFF and right for ON condition. For both groups, the electrodes in the central area have a higher level of node degree than that of other regions. Color bar indicates the magnitude of node degree. B. Spatial difference pattern for comparison of node degree between two conditions (OFF vs. ON). The labeled channels show the identified significant negative cluster (OFF < ON, p = 0.0140) using cluster-based permutation test. Color bar indicates the statistical T-value.

### 4.5. No significant change in the global network topology: either in network segregation or network integration measure

To answer the question whether the global network structure is modulated by medication, we estimated the two fundamental features of a network: the global efficiency for measuring functional network integration and the clustering coefficient for measuring network functional segregation. We report the comparison results for both of the measures across a wide range of proportional thresholding values (36% to 4%, with a step of 2%) between the two conditions. Since it has been shown that differences in overall functional connectivity could have predictable consequences for between-group differences in network topology (van den Heuvel et al., 2017), we here first checked whether in our data there could be a possible bias for the comparison. However, no significant difference in overall FC between condition comparisons was found (Wilcoxon signed rank test, two tailed, p = 0.1514). Thus, the overall FC is probably not a significant bias in the comparisons we performed as shown below. As seen in Figure 5A, across the whole range of thresholding (36%–4%), the mean GE across subjects in the OFF condition (in black) almost overlapped with that from the ON condition (in red). As for clustering coefficient, the grand mean of CC in the OFF condition (black line) showed higher values than those in the ON condition (red line) across all thresholding values (Fig. 5B). However, the statistical comparison did not indicate a significant difference in global efficiency (p > 0.05, p values shown in dashed orange line, right y axis), or in clustering coefficient between the two conditions (p> 0.05, p values shown in dashed orange line, right y axis). Thus, controlling for the overall FC values and across a wide range of thresholding values, we were not able to demonstrate a significant impact of medication on global network configuration.

**Figure 5.**
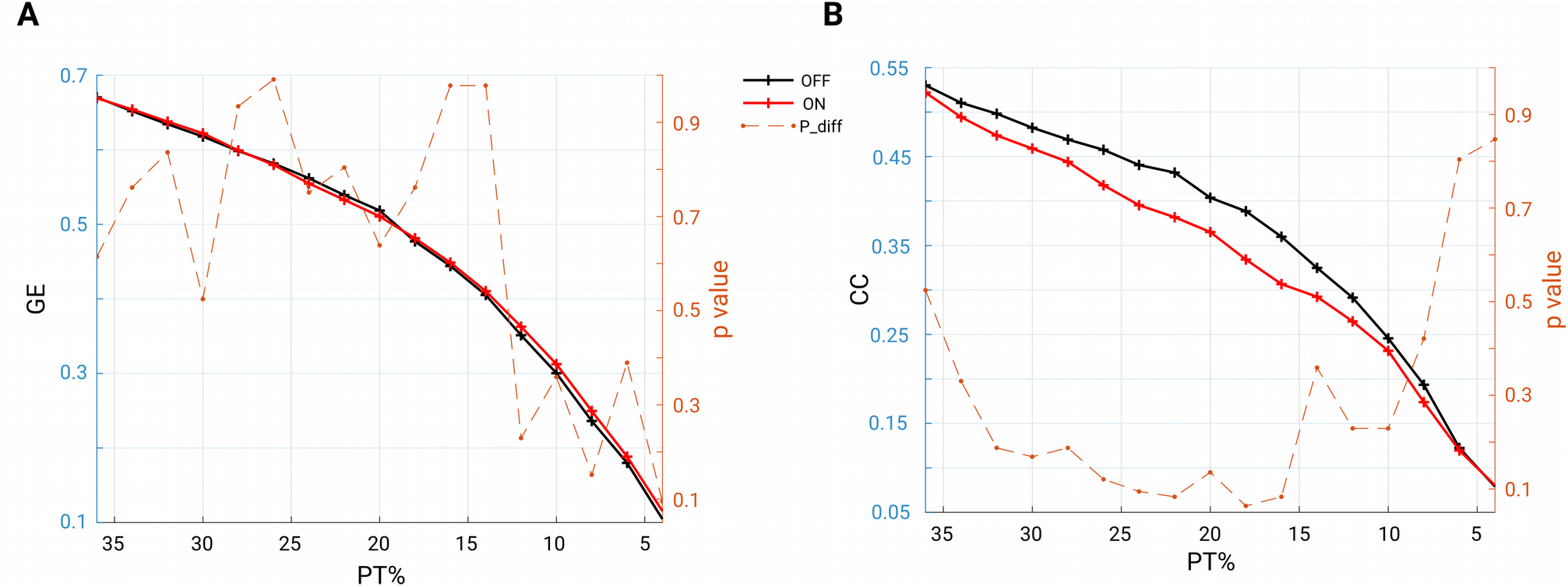
A. Mean GE estimated from beta band functional connectivity across subjects within each group (OFF in black and ON in red) across a wide range of thresholding values (36% to 4% in step of 2%). The dashed orange line represents the p values (right y axis) for the comparisons: none are below 0.05. B. Same analysis, but for CC: across a wide range of thresholding values no significant difference was observed between the conditions (OFF vs. ON).

### 4.6. Spectral slope (local and global) predicts the network global efficiency in OFF medication

We investigated a possible relationship between spectral slope and network topology. First, we averaged the spectral slope across all channels to represent an overall slope (referred to as global slope) for each subject. Spearman’s correlation was performed between global slope and network metrics (GE and CC) derived under an exemplary thresholding value at 20% in both groups. As shown in the scatter plot in Figure 6A, global efficiency negatively correlated with global slope (Rho = -0.7643, p < 0.001) in the OFF condition. In contrast, no such association was observed in the ON condition (Rho = - 0.1036, p = 0.7144). Next, we performed a correlation analysis for the channel-wise slope (referred to as local slope) and network global efficiency in the OFF condition. This analysis revealed a significant negative relationship between local slope values and network global efficiency as shown in the topographical map (channels demonstrating significance are in blue, FDR-corrected) in Figure 6B, and this relationship was most pronounced in the left centro-parietal area. There was no significant relationship between local slopes and GE in the ON condition. In addition, we examined if the relationship we observed at the 20% thresholding could be obtained regardless of the specific thresholding value. We performed the correlation analyses between global slope and network GE across the whole range of thresholding values (36%–4% with a step of 2%) in the OFF group. As shown in Figure 6C, almost across all PT%, the negative association between global slope and network GE was present consistently (p < 0.05, p values shown in dashed orange line, right y axis), except under an extreme thresholding value of 4%. The spatial correlation pattern between local slope and network GE was also examined under the same range of thresholding values, and consistently negative relations between local slope from the centro-parietal region and network GE were observed (see supplementary material). These results showed that global slope negatively correlated with network global efficiency across a wide range of thresholding values, and a further topographical correlation map between local slope and network GE demonstrated a region-specific pattern.

**Figure 6.**
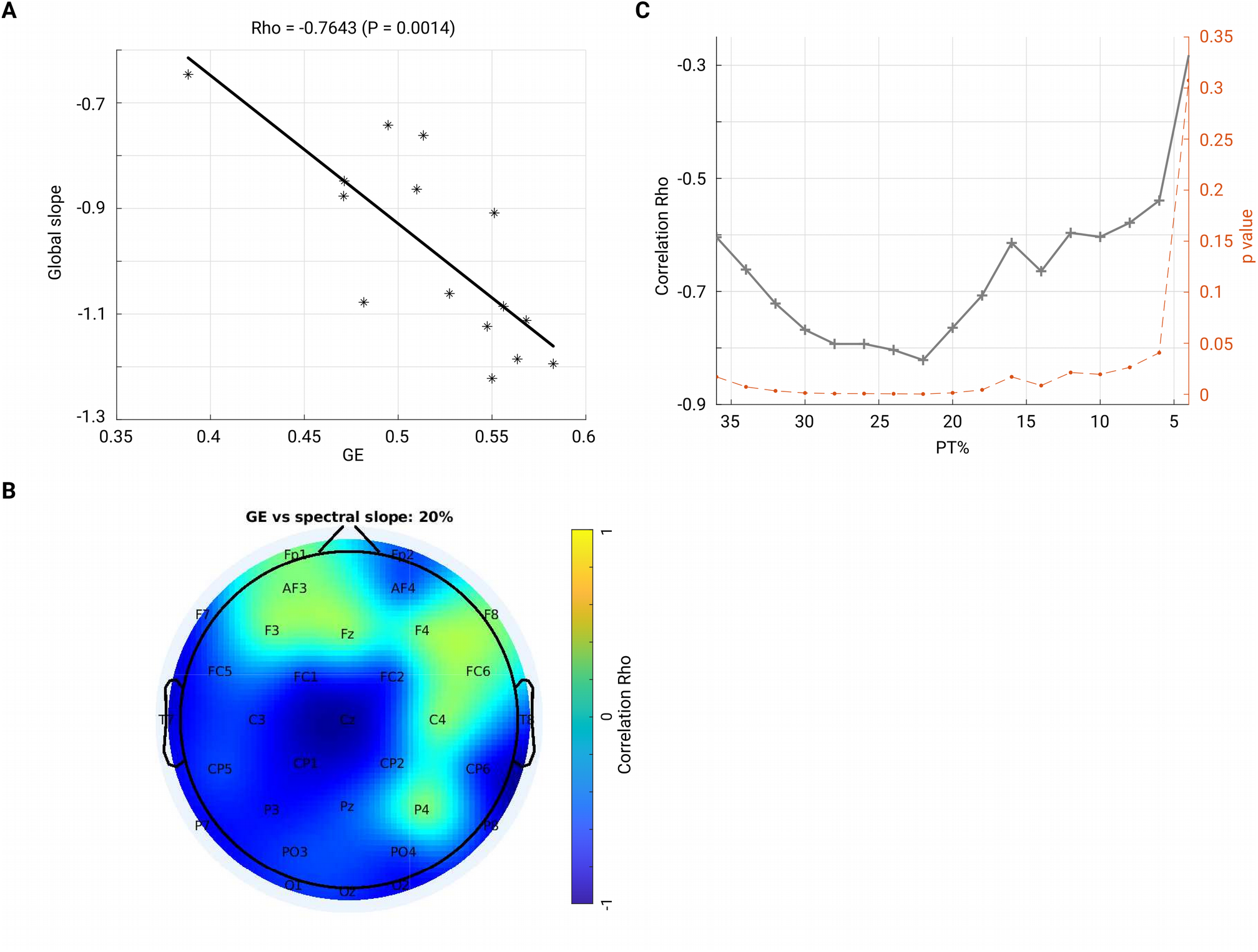
A. Scatter plot shows that global efficiency for the beta band network (under thresholding value 20%) negatively correlates with the global slope (averaged slope across the whole head) in the OFF condition. Each dot represents one subject. B. Spatial pattern for the correlation between local slope and global efficiency (beta network and under 20% thresholding). The channels in blue indicate significant correlation after FDR correction. Color bar indicates the correlation coefficient value. C. Through a family of thresholding values (36%–6%, with a step of 2%), there was a significantly negative relationship (p < 0.05, p values shown in dashed orange line, right y axis) between global efficiency and global slope.

## 5. Discussion

In this study, we investigated local and global changes induced by dopaminergic medication in a cohort of PD patients using non-oscillatory spectral slope measure and connectivity analysis in resting state EEG. Locally, we estimated the slope of the non-oscillatory wideband background activity and showed that the left central region had a significantly decreased (steeper) spectral slope during the ON compared to OFF medication state. In addition, in ON compared to OFF, we observed an increase in the functional connectivity in the beta band, mainly between centro-parietal and frontal regions. Further, graph theory-based analysis showed an enhanced node centrality in particular in the centro-parietal regions but no significant alteration in the complex level of network topology (global efficiency or clustering coefficient). Lastly, we found a strong negative relationship between spectral slope (locally and globally) and network’s global efficiency in the OFF condition, where a flatter slope was associated with a smaller degree of global efficiency of the functional network. These findings provide further evidence for the engagement of multiple cortical regions in response to dopaminergic medication in PD, which in turn may indicate that the therapeutic efficacy of dopaminergic medication may relate to both regional and global changes in cortical activity.

### 5.1. Non-oscillatory background spectral slope

Using multi-channel resting state EEG, we observed that patients with PD in the medication OFF condition had an increased (flatter) spectral slope compared to medication ON condition. This effect was found to be spatially specific to the left central region. The spectral slope, a metric to quantify this background power spectrum, has been reported to be altered in the first year of development, healthy aging and in mental disorder such as schizophrenia (Donoghue et al., 2020; Molina et al., 2020; Peterson et al., 2017; Schaworonkow & Voytek, 2021), and could also predict the dynamic behavioural outcome in working memory tasks (Donoghue et al., 2020; Voytek et al., 2015). In our study, we observed that the spectral slope steepened in ON compared to OFF conditions, and we assume that patients in the OFF condition had flattened slopes compared to healthy controls, and dopaminergic medication could normalize this flattening effect. This assumption is well aligned with previous studies demonstrating that healthy aging is accompanied by flattening of the spectral slope (Cesnaite et al., 2021; Voytek et al., 2015), and neural electrophysiological biomarkers associated with PD are already present in the apparently healthy aging brain (Zhang et al., 2021). The effect was found most pronounced in the left central area (strongest at C3 electrode in the detected cluster), which might indicate a modification over the sensorimotor area by the medication. The broadband spectral slope underlying the dopamine medication modulation effect in patients with PD may thus potentially serve as a biomarker sensitive to dopamine replacement therapy. At the same time, even though we carefully cleaned the data and removed artifacts which might contribute to the estimation of spectral slope, we could not completely rule out this confounder. However, we would like to emphasize that this is unlikely to drive the effect of spectral slope we observed, otherwise one would expect a spatial pattern which shows strongest difference over the frontal or temporal areas (which cover large muscle groups and prone to be contaminated by the muscle activity). Additionally, as we mentioned before, the spectral slope has been shown to index the E/I balance, and we will discuss the implication of this finding below (Discussion section *Spectral slope and network global efficiency: local E/I balance and global network*).

### 5.2. Power of beta oscillation

Previous studies have demonstrated an increase in cortical beta-band power in PD compared to healthy controls and alleviated beta band synchrony after medication administration (Stanzione et al., 1996) and attenuation by deep brain stimulation (Whitmer et al., 2012). On the other hand, other studies have also reported an opposite effect — an increase of beta band power after dopaminergic medication (Melgari et al., 2014). In addition, some studies demonstrated that dopaminergic medication did not have any effect on cortical beta power (George et al., 2013; Miller et al., 2019; D. Stoffers et al., 2007; Swann et al., 2015). Importantly, all previous PD studies on this topic have only considered total power of beta without separating it into oscillatory and 1/f aperiodic components. In the present study, we tested the impact of the removal of the aperiodic part of the spectrum on the estimation of oscillatory power. We found that a conventional approach to estimate oscillatory power based on the raw PSD resulted in a non-significant difference in beta band in the PD OFF compared to ON state. After accounting for the spectral slope changes, a marginal increase of beta power was detected in the centro-parietal regions in the comparison between the ON and OFF condition, yet this difference failed to reach significance. Our data thus suggests that even though the beta-band power estimation by the conventional approach might be partly affected by the background wideband PSD spectra, correcting the effect still does not yield a clear and statistically significant difference between the ON and OFF conditions. Thus, in line with some previous studies (George et al., 2013; Miller et al., 2019; Swann et al., 2015), we further confirm that with and without considering the background slope effect, there was no difference in beta power between the medication conditions. Yet, we suggest that future studies should take into account the effect of the aperiodic spectral component for the comprehensive evaluation of oscillatory power changes in PD.

### 5.3. Functional connectivity

We observed a significant increase in functional connectivity of beta oscillations in the ON compared to OFF condition, in particular between the centro-parietal regions with frontal regions. Previous studies have demonstrated a presence of beta-band coherence between STN (subthalamic nucleus) and multiple cortical regions, including sensorimotor (Hirschmann et al., 2011, 2013; Litvak et al., 2011), parietal and frontal areas (Litvak et al., 2011) in the OFF medication condition in patients with PD. Dopaminergic medication can also alter the beta-band connectivity between STN and cortical regions (Hirschmann et al., 2013; Litvak et al., 2011; Diederick Stoffers et al., 2008; van Wijk et al., 2016). As for the cortico-cortical connectivity, dopaminergic medication administration was shown to either reduce interactions between cortical areas (George et al., 2013; Heinrichs-Graham et al., 2014; Pollok et al., 2013; Silberstein et al., 2005) or not to produce any significant changes (Miller et al., 2019). In a very recent study using combined STN-LFP (local field potential) and MEG recordings, the authors discovered differential effects of dopaminergic medication in different levels of networks (Sharma et al., 2021). Specifically, in the cortico-cortical network, sensorimotor-cortical connectivity across multiple regions was enhanced in the beta band during the ON medication state. Therefore, our observations of the enhancement of such a coherent fronto-parietal motor network in the ON condition is consistent with this recent report. Such enhancement of functional connectivity is partially in agreement with another study which employed simultaneous fMRI/EEG recordings and showed that a higher dose of dopaminergic medication increased functional connectivity between motor areas and the default mode network in fMRI, whereas EEG connectivity remained unaffected (Evangelisti et al., 2019). In general, the dopaminergic effect over the cortico-cortical motor network might relate to the motor decision-making associated network, which has been shown to involve cortical fronto-parietal regions (Siegel et al., 2015), or it might relate to the default-mode network changes associated with non-motor symptoms in PD as suggested by other fMRI studies (L. lin Gao & Wu, 2016). Notably, a recent EEG study in PD using source localization demonstrated the presence of strong phase-amplitude coupling between the phase of beta and the amplitude of broadband gamma oscillations in a variety of cortical regions (including sensorimotor, somatosensory, and prefrontal areas) involved in motor and executive control (Gong et al., 2021). In line with this study, our findings of increased connectivity between centroparietal-frontal regions after dopaminergic medication further emphasize the importance of cortico-cortical connections in PD. These electrophysiological findings are consistent with previous fMRI studies suggesting a critical role of motor circuitry in PD in response to dopamine administration (Shen et al., 2020).

### 5.4. Global and local network organization

Using graph theory, we demonstrated that in the ON condition, there was a significant increase in node degree in centro-parietal regions implying that these regions became more influential in the communication within the network. However, the network topology does not seem to undergo a major re-configuration as we did not identify significant changes in global efficiency or clustering coefficients in the brain network. This seems consistent with findings of previous studies in which PD patients were compared to healthy controls and no differences in topographical properties were found at the global level either in fMRI (Ruan et al., 2020) or in EEG in all frequency bands (Hassan et al., 2017). Another previous study also investigated the topographical structure of functional network using graph analysis based on MEG of patients with PD (Olde Dubbelink et al., 2013). Compared to healthy controls, their longitudinal study revealed a tendency toward a more random brain functional organization which was associated with lower local integration in multiple frequency bands and lower global efficiency in the upper alpha band. However, another study using EEG found an increase in local integration and a decrease in global efficiency across all the frequency bands in PD compared to healthy subjects (Utianski et al., 2016). In the present study, we explored the alterations in a functional spectral network using graph metrics and showed that dopaminergic medication intake did not significantly alter the brain network organization but did exert a significant enhancement in node degree of some particular regions within the network. The absence of significant changes in global integration and segregation of the functional network might suggest that dopaminergic medication does not re-configure the network at a global organizational level. Instead, these observations appear to imply that the brain network as a whole does not respond to medication at the complex (global integration and segregation) but rather at the low-level network topology (local node). It would be interesting for future studies to test whether this relates to the clinical improvement of symptoms and whether it is possible to significantly alter the network organization through different therapeutic interventions based on brain stimulation.

### 5.5. Spectral slope and network global efficiency: local E/I balance and global network

A steeper spectral slope after dopaminergic medication intake was evident in PD. Interestingly, the wide-band background spectral slope has recently been shown to be linked to the excitation and inhibition balance (E/I balance) of the recorded site both in simulations and experimental data, demonstrating that a steeper slope correlates with a state characterized by a more inhibitory compared to excitatory tone (Colombo et al., 2019; R.Gao et al., 2017). Following the E/I balance hypothesis of the spectral slope, a steeper slope after medication, observed in this study, may indicate that dopamine induced a state characterized by stronger inhibition over excitation. This line of interpretation agrees with previous TMS (transcranial magnetic stimulation) studies reporting a reduction of intracortical inhibition at rest in PD OFF medication (Cantello, 2002; Hanajima et al., 1996; Ridding et al., 1995) and an enhancement of evoked inhibitory activity (reflected in late TMS-evoked activity and beta TMS-evoked oscillations) after dopaminergic medication intake (Casula et al., 2017).

In addition, we found a close relationship between broadband non-oscillatory background activity measured by the spectral slope and the beta-band global efficiency of the functional network. Global network efficiency represents the ability of integration of activity of widely distributed regions within a network, impacting information transmission and communication (Bullmore & Sporns, 2012). Notably, a previous simulation work demonstrated that synaptic E/I balance is crucial for efficient neural coding (S. Zhou & Yu, 2018), and the local E/I ratio plays a role in information transmission at large scale brain level (Deco et al., 2014). This theory concurs with our findings: the local and global spectral slope, reflecting the local and global tune of E/I balance, is closely associated with the functional network global integration property. The negative relationship between them implies that more excitation over inhibition corresponds to a lower level of functional network integration. Consistently, a recent study from both fMRI recording and simulation data showed that the local E/I ratio could have a significant impact on the organization of whole brain functional networks: global efficiency of the functional network is an inverted-U shaped function of local E/I ratio and the more deviation from the balanced E/I state (in either direction), the lower global efficiency of the whole functional network (X. Zhou et al., 2021). Our observation about the relationship between local and global slopes with the global network integration property can potentially be explained by this model: in OFF medication, an imbalanced E/I state (indexed by flatter slope) deviating from balanced E/I ratio exerts a monotonous negative relation with functional network GE. Such a close association did not hold for the medication ON group. We assume that the medication moves the network back closer to a more balanced state, reflected in a steeper spectral slope (steepening of the flattened slope in OFF state); thus, functional network organization was no longer closely related to the E/I, since in a close-to balanced E/I state the global efficiency would rather remain stable (i.e., it reaches a maximum at the optimal E/I state).

The spatial distribution of local slope and global efficiency demonstrated a specific pattern where the slope from the centro-parietal regions showed strongest relations with the GE of the brain network. In line with previous fMRI studies demonstrating that the nodal property of the parietal cortex is closely associated with motor outcome and decreased with progressing disease stage (Hoehn and Yahr stage) in PD (Fang et al., 2017; Sang et al., 2015; Suo et al., 2017), we assume that centro-parietal regions play an important role in orchestrating the whole global network organization. This is congruent with the finding that the connectivity patterns in these cortical regions are also affected by dopaminergic medication, as discussed above.

## 6. Limitations

The first limitation of this study is that due to a rather low density of electrodes, we performed all connectivity analysis in sensor space. Thus, we refrain from making any conclusions about the specific structure of the networks (e.g., small-world, scale-free networks) as is also suggested in a critical study on the application of graph measures in EEG/MEG (Kaminski & Blinowska, 2018). It should also be noted that even if the analysis were to be conducted in source space, the volume conduction issue may still be present. Importantly, we applied a connectivity measure that is specifically used to overcome the volume conduction issue. Moreover, we were able to show that our findings remained consistent for a wide range of thresholds for the networks’ properties.

Another limitation of our study is that clinical measures were not available and therefore, we could not associate EEG measures with the severity of clinical symptoms. We acknowledge this and suggest that future studies could include such a design so that the link between EEG parameters and clinical phenotypes can be explored. Future work should test whether a n d h o w local and global EEG parameters relate to clinical symptoms.

## 7. Conclusion

Using multi-channel resting EEG recordings in PD patients, we showed differential effects of dopaminergic medication on local non-oscillatory components and connectivity parameters. Both from the local-level and brain-network perspective, the centro-parietal area was identified as the region where significant alterations in non-oscillatory wideband activity, measured by spectral slope and node centrality within the spectral functional network in the beta band, occurred. However, the network’s global topologies, namely global integration (measured by global efficiency) and global segregation (measured by clustering coefficient) remained unaffected by the dopaminergic medication. Furthermore, during the OFF state, a close association between the spectral slopes (local and global) and network global integration was observed. These findings align with the theory that local E/I balance impacts global network structure, which might in turn demonstrate a crucial role of local non-oscillatory dynamics in forming the functional global integration in PD.

## Data Availability

All the data in this study are available online at https://openneuro.org/datasets/ds002778.

## Acknowledgements

We thank Tilman Stephani for valuable discussions on the manuscript. This work was supported by Deutsche Forschungsgemeinschaft (German Research Foundation) (Project ID: 424778381 TRR 295).

## Supplemental material

1. Spatial correlation patterns between local slope and beta band network GE across a wide range of thresholding values. Spatial specificity over the centro-parietal region is generally consistent across a family of thresholding values (36%–18%). At the PT% of 14%, a significant negative relationship was still present over channel Cz. For the higher PT% values (12%–4%), no significant relationship remained after multiple testing correction.

**Figure S1.**
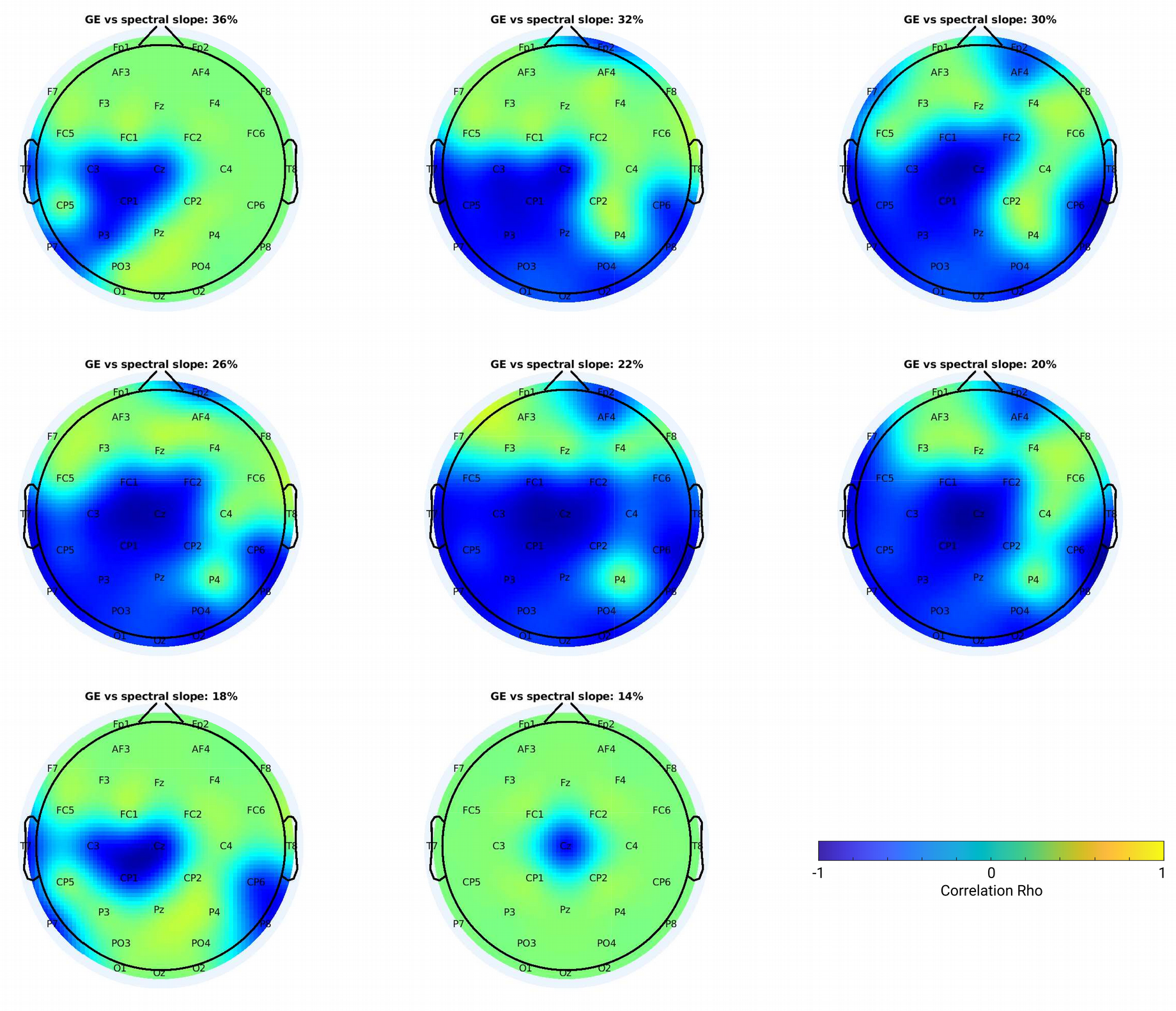
Spatial patterns for the correlation between local slope and global efficiency (beta network and under a variety of thresholding values). Significant channels are shown in blue (p < 0.05) after FDR correction. Color bar indicates the correlation coefficient value.

